# Polygenic score for body mass index is associated with disordered eating in a general population cohort

**DOI:** 10.1101/2020.04.03.20051896

**Authors:** Mohamed Abdulkadir, Moritz Herle, Bianca De Stavola, Christopher Hübel, Diana Santos Ferreira, Ruth J F Loos, Rachel Bryant-Waugh, Cynthia M. Bulik, Nadia Micali

## Abstract

**Background:** Disordered eating (DE) is common and is associated with body mass index (BMI). We aimed to investigate if genetic variants for BMI were associated with DE.

**Methods:** BMI polygenic scores (PGS) were calculated for participants of the Avon Longitudinal Study of Parents and Children (ALSPAC; N= 8,654) and their association with DE tested. Data on DE behaviors (e.g. binge eating, and compensatory behaviors) were collected at ages 14, 16, 18 years, and DE cognitions (e.g. body dissatisfaction) at 14 years. Mediation analyses determined whether BMI mediated the association between the BMI-PGS and DE.

**Results:** The BMI-PGS was positively associated with fasting (OR= 1.42, 95% CI=1.25, 1.61), binge eating (OR=1.28, 95% CI= 1.12, 1.46), purging (OR= 1.20, 95% CI= 1.02, 1.42), body dissatisfaction (Beta= 0.99, 95% CI= 0.77, 1.22), restrained eating (Beta=0.14, 95% CI= 0.10, 1.17), emotional eating (Beta= 0.21, 95% CI= 0.052, 0.38), and negatively associated with thin ideal internalization (Beta= −0.15, 95% CI= −0.23, −0.07) and external eating (Beta= −0.19, 95% CI= −0.30, −0.09). These associations were mainly mediated by BMI.

**Conclusions:** Genetic variants associated with BMI are also associated with DE. This association was mediated through BMI suggesting that weight potentially sits on the pathway from genetic liability to DE.

## Introduction

Disordered eating (DE) behaviors including fasting, binge eating, and related cognitions (such as body dissatisfaction) are widely prevalent in the general population (14-22%) and are considered behavioral and psychological features of a clinical diagnosis of anorexia nervosa (AN), bulimia nervosa (BN), and binge-eating disorder (BED) [1–5]. DE typically arises during pre-adolescence and adolescence [3,5,6] and individuals with DE are at greater risk for mood disorders, psychosocial impairment, and suicidal behavior, as well as at elevated risk for developing a full eating disorder [7,8]. Identification of risk factors that may contribute to DE is an active area of inquiry and may offer new prevention and therapeutic interventions as the current treatment strategies for eating disorders (ED) are limited in their efficacy [9,10].

DE and related cognitions are moderate to highly heritable with estimates from twin studies ranging between 20-85% making it suitable for genetic analyses [11,12]. Polygenic score (PGS) analyses are an effective method that allow for direct testing of whether common genetic variants associated with one trait are also associated with another [13,14]. PGS approaches to understanding DE are of interest as they allow to interrogate the share genetic etiology of two traits that are associated at the phenotypical level, such as DE and BMI [15,16]. Previous research has highlighted that DE and related cognitions are present across the entire weight spectrum, including AN (at one extreme of the weight spectrum) and BED (often at the other extreme of the weight spectrum)[17]. In addition, children who develop an ED have been shown to follow different BMI trajectories prior to diagnosis [18–20]. More specifically, children who go on to develop AN, show consistently lower childhood BMI, whereas children who later develop BED show higher premorbid childhood BMIs [18].

It is well established that BMI has a substantial genetic component [21] with the SNP-based heritability ranging between 17-27% [22]. Recent studies have shown a negative genetic correlation between AN and BMI (r_g_ ∼ −0.24) suggesting shared genetic etiology between AN and BMI, whereby genetic variants associated with higher BMI were associated with lower risk for AN [23,24] However, less is known about the extent to which DE shares genetic etiology with BMI.

The first study to investigate the association between DE and a BMI-PGS found that a BMI-PGS is associated with weight loss behaviors [25]. Nagata et al. found that the BMI-PGS was associated with a higher odds of weight loss behaviors (e.g., dieting, vomiting) and a lower odds of weight gain behaviors (e.g., eating more or different foods than normal). In addition, Nagata et al. found that the association between the BMI-PGS and weight loss behaviors was mediated by measured BMI; a higher BMI-PGS was associated with a higher measured BMI which in turn was associated with a higher odd of engaging in weight loss behaviors. However, this study [25] investigated this association only in a sample of young adults and evidence is currently lacking in adolescence; a critical period for the development of ED and DE. Furthermore, this study failed to include pre-morbid BMI (measurement of BMI prior to DE) into the mediation analyses which complicates establishing potential causal pathways that lead from measured BMI to DE.

In the current study, we investigated whether a BMI-PGS is longitudinally associated with a broad range of DE behaviors and cognitions in a large UK-based population study across adolescence. We hypothesized that the BMI-PGS will be positively correlated with binge eating, emotional eating, and inappropriate compensatory behaviors, such as purging and fasting for weight loss. Based on a previous positive correlation of higher body weight with higher adolescent body dissatisfaction [3] and restrained eating [25,26], we hypothesized that the BMI-PGS will positively correlate with later body dissatisfaction and restrained eating. We also expected a negative association between BMI-PGS and both thin ideal internalization and high external eating—both of which have been found to be negatively correlated with weight or BMI in population-based studies[26,27]. One possible pathway that genetic risk could lead to DE could be through changes in measured BMI which then could lead to increased risk for DE; therefore, we aimed to determine the role of measured BMI as a mediator of these associations. Considering the prospective nature of the data, we also examined developmental differences, determining whether the strength of the association between the BMI-PGS and DE differ across the three time-points during adolescence.

## Methods and Materials

### Participants

The ALSPAC study is an ongoing population-based birth cohort study of 15,454 mothers and their children residing in the south west of England (UK) [28–31]. Participants are assessed at regular intervals from birth, using clinical interviews, self-report questionnaires, medical records, and physical examinations. The study website contains details of available data through a fully searchable data dictionary: http://www.bristol.ac.uk/alspac/researchers/our-data/. One sibling per set of multiple births was randomly selected to guarantee independence of participants (n = 75). Furthermore, individuals that are closely related to each other defined as a phi hat > 0.2 (calculated using PLINK v1.90b) were removed; this meant removal of any duplicates or monozygotic twins, first-degree relatives (i.e., parent-offspring and full siblings), and second-degree relatives (i.e., half-siblings, uncles, aunts, grandparents, and double cousins). Ethical approval for the study was obtained from the ALSPAC Ethics and Law Committee and the Local Research Ethics Committees. Informed consent for the use of data collected via questionnaires and clinics was obtained from participants following the recommendations of the ALSPAC Ethics and Law Committee at the time.

### Measures

#### Binary outcomes

Information on fasting, binge eating, and purging, were assessed at ages 14, 16, and 18 years using questions modified from the Youth Risk Behavior Surveillance System questionnaire [32]. For fasting (N age 14 = 4,584, N age 16 = 3,844, N age 18 = 2,586), participants were asked “During the past year, how often did you fast (not eat for at least a day) to lose weight or avoid gaining weight?” with the response options “Never”, “Less than once a month”, “1-3 times a month”, “once a week”, and “2 or more times a week”. This variable was dichotomized as fasting at least once a month in the previous year versus no fasting [6]. For binge eating (N age 14 = 4,144, N age 16 = 3,336, N age 18 = 1,910), participants were asked how often they engaged in overeating (eating a very large amount of food) in the previous year. Participants who answered positively to this question were subsequently asked whether they felt out of control during these episodes. We dichotomized the binge eating variable as eating a very large of amount of food at least once a month (with the feeling of loss of control) versus no binge eating. Regarding purging (N age 14 = 4,588, N age 16 = 3,871, N age 18 = 2,582), participants were asked how often they self-induced vomiting or had taken laxatives (or other weight loss medications) to lose weight or avoid weight gain in the previous year; this variable was then dichotomized as purging at least once a month versus no purging.

#### Continuous outcomes

All continuous DE outcomes were assessed at age 14 years and included thin ideal internalization, body dissatisfaction, emotional eating, external eating, and restrained eating. Thin ideal internalization (N = 4,496) was assessed using the Ideal-Body Stereotype Scale-Revised with gender-specific items used to assess different aspects of appearance-ideal internalization for boys (six items) and girls (five items) [6,33,34]. The responses were rated on a five-level Likert scale from “strongly agree” to “strongly disagree” and the items from this scale were summed to obtain a total score; a higher score corresponded with increased internalization of the thin ideal Body dissatisfaction (N = 4,625) was assessed using the Body Dissatisfaction Scale [34,35]. Participants were asked gender-specific questions to rate their satisfaction with nine body parts with responses on a six-level Likert scale ranging from ‘extremely satisfied’ to ‘extremely dissatisfied’. From this scale a continuous score was constructed in which a higher score corresponded with a higher body dissatisfaction [6,34]. Restrained eating (N = 4,530), emotional eating (N = 4,345), and external eating (N = 3,995) were assessed using a modified version of the Dutch Eating Behavior Questionnaire (DEBQ; 22). The restrained eating subscale was assessed with 2 questions, 14 for the emotional eating subscale, and 7 for external eating [34,37].

#### Body Mass Index

BMI (weight/height^2^) was calculated using objectively measured weight and height during a face to face assessment at age 11 years (N = 5,902) [28–30]. Height was measured to the nearest millimeter using a Harpenden Stadiometer (Holtain Ltd.) and weight was measured using the Tanita Body Fat analyzer (Tanita TBF UK Ltd.) to the nearest 50 grams. Age- and sex-standardized BMI z-scores (zBMI) were calculated according to UK reference data, indicating the degree to which a child is heavier (>0) or lighter than expected according to his/her age and sex [38].

### Genotyping

Genotype data were available for 9,915 children out of the total of 15,247 ALSPAC participants. Participants were genome-wide genotyped on the Illumina HumanHap550 quad chip. Details of the quality control checks are described in the Supplementary note.

### PGS calculations

PGS were derived from summary statistics of the Genetic Investigation of Anthropometric Traits (GIANT) consortium, referred to as the discovery cohort [39]. The calculation, application, and evaluation of the PGS was carried out with PRSice (2.1.3 beta; github.com/choishingwan/PRSice/) [40,41]. PRSice relies on PLINK to carry out necessary cleaning steps prior to PGS calculation [40,42]. Strand-ambiguous SNPs were removed prior to the scoring. A total of 1,488,001 SNPs were present in both the discovery and in the target cohort. Clumping was applied to extract independent SNPs according to linkage disequilibrium and *P*-value: the SNP with the smallest *P*-value in each 250 kilobase window was retained and all those in linkage disequilibrium (r2 > 0.1) with this SNP were removed. To calculate the PGS, for each participant the sum of the risk alleles was taken and was then weighted by the effect size estimated from the discovery cohort. The PGS were calculated using the high-resolution scoring (PGS calculated across a large number of P-value thresholds) option in PRSice, to identify an optimal p value threshold at which the PGS is optimally associated with the outcome.

### Statistical analyses

#### Sensitivity analyses (Linkage Disequilibrium Score Regression)

The discovery GWAS from which the BMI-PGS was derived included participants from the UK Biobank [39]. Considering a potential overlap in participants between ALSPAC and the UK Biobank we performed a BMI GWAS with biological sex as a covariate using PLINK in the ALSPAC sample [42]. Subsequently we determined the genetic covariance with the BMI GWAS [39] using linkage disequilibrium score regression (LDSC) v1.0.0. [43].

#### Regression analyses

Logistic regressions models were applied for binary outcomes (e.g., fasting at age 14, 16, and 18 years) and linear regression models were used for continuous variables (e.g., thin ideal internalization) with biological sex and the first four ancestry-informative principal components as covariate in all models. We have also tested for an interaction between the BMI-PGS and sex and have conducted gender stratified analyses.

As confirmatory analyses, we investigated the association between the BMI-PGS and BMI at age 11 and 18 years and the association between the BMI-PGS and the age and sex standardized BMI (i.e., BMI z-scores) measured at age 11 years. We then investigated the association of DE symptoms at age 14 years, except for purging due to its low endorsement and hence exclusion from our analyses (N purging at age 14 years = 74). We repeated the same analyses at age 16 and 18 years for fasting, binge eating, and purging. We report the regression models which explain the largest R^2^ or Nagelkerke’s pseudo-R^2^ at the optimal BMI-PGS P-value threshold. For the binary outcome measures we report the Nagelkerke’s pseudo-R^2^ on the liability scale [44]. Empirical P-values were calculated through permutation (N = 11,000) that account for multiple testing (i.e., number of P-value thresholds tested) and overfitting [40]. We additionally calculated False Discovery Rate-corrected Q-values adjusting for the number of phenotypes tested [45]. The significance threshold was met if the False Discovery Rate-adjusted Q was < 0.05.

#### Generalized linear mixed models (GLMM)

To understand whether the association of the BMI-PGS with DE differs across the ages (i.e., developmental differences), we used generalized linear mixed models, including BMI-PGS, biological sex, the first four ancestry-informative principal components as fixed effects, and age at self-report as an interaction term. We included random intercepts for each individual in the model, to take into account variance in DE that is due to inter-individual differences. To generate comparable results across the ages, we calculated the PGS at a P-value threshold (P_t_) of 1 for each behavioral trait at each age.

#### Exploratory causal mediation analysis

The association between BMI-PGS and adolescent DE could to a certain extent be explained through childhood BMI prior to assessment of DE. We used sex and age adjusted BMI z-scores (zBMI) measured around the age of 11 years (prior to assessment of DE and related cognitions) to capture a possible pathway linking genetic liability, measured by the standardized BMI-PGS to DE (Figure 1). We estimated the BMI-PGS effects that are mediated and not mediated by childhood zBMI scores using concepts proposed in modern causal inference; the natural direct and indirect effects [46]. The natural direct effect (also known as the “average direct effect”, ADE) measures the expected risk difference of the binary outcome measures (e.g., fasting) had the BMI-PGS been hypothetically set to change by 1 SD from 0 to 1, while at the same time childhood zBMI scores had been set to take its natural value (i.e., the value that would be experienced had the BMI-PGS been set at the reference value of 0). For the continuous outcome measures (e.g., thin ideal internalization), the ADE measured as the mean difference of the outcome had the BMI-PGS been set to change by 1 SD while childhood zBMI scores was set to take its natural value. The natural indirect effect (“average causal mediated effect”, ACME) measures the expected risk difference in binary outcome measures had the BMI-PGS been hypothetically set to take the value 1, while at the same time childhood zBMI scores had been set to take its potential values had BMI-PGS been set to 0 or 1. The ADE was estimated in a similar fashion for the continuous outcome measures except that the ACME measured the expected mean difference of the outcome. When summed together, these direct and indirect effects give the total causal effects and therefore are useful measures of the contribution made by a particular pathway to a causal relationship. We estimated these effects, expressed as risk differences/mean differences, using the R package ‘mediation’ (version 4.4.6; 32). Our analyses were controlled for biological sex, and the first four ancestry-informative principal components. Furthermore, we assumed that there were no additional unaccounted confounders nor any intermediate confounders [46].

**Figure 1:**
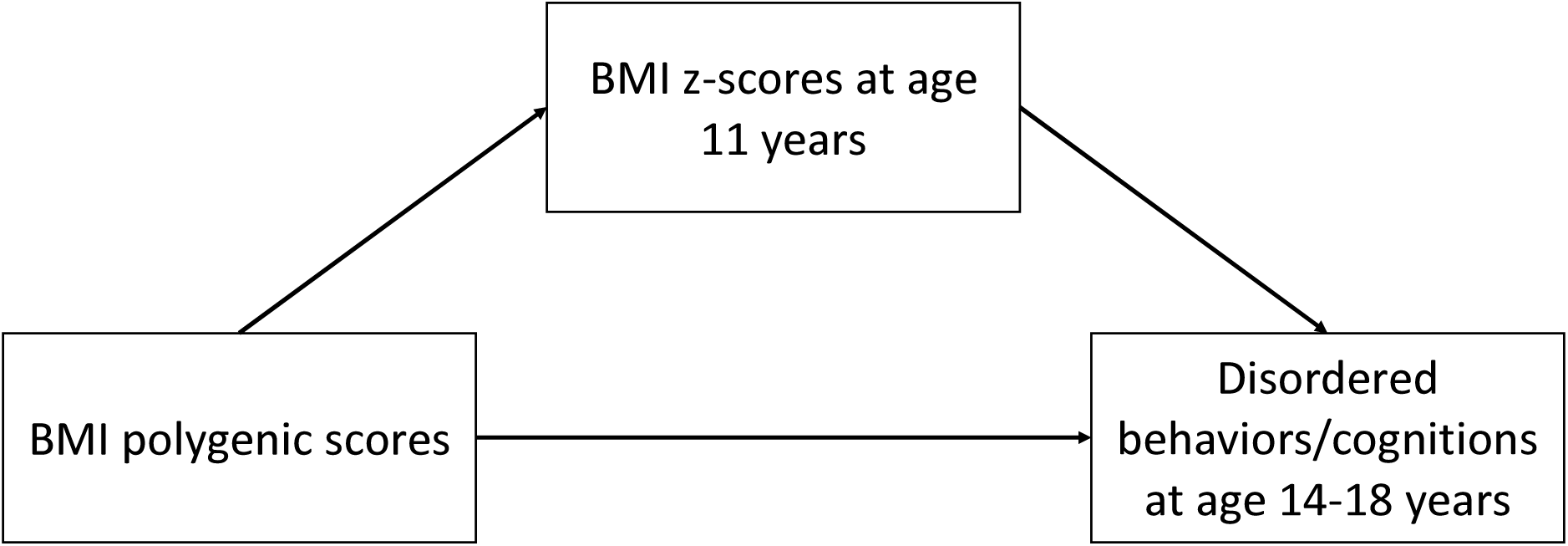
Directed acyclic graph of the causal mediation analyses to understand the role of body mass index z-scores (zBMI) at age 11 years in the association between the BMI polygenic scores (PGS) and disordered eating behaviors and cognitions during adolescence. The BMI-PGS was derived from summary statistics of the genome-wide association study (GWAS) carried out by the Genetic Investigation of Anthropometric Traits (GIANT) consortium [39] and were calculated for participants in the Avon Longitudinal Study of Parents and Children [28–31]. The sex and age adjusted BMI z-scores were included as a mediator in the causal mediation analyses that was carried out using the R package ‘mediation’ (version 4.4.6; 32). The ‘mediation’ package is based on concepts proposed in modern causal inference [46]. Prior to the mediation analyses the BMI-PGS was standardized (to mean zero and standard deviation of one) and the analyses were controlled for biological sex and the first four ancestry-informative principal components.

#### Missing data

Considering the longitudinal nature of ALSPAC, attrition is unavoidable and the data for analyses was missing at random (MAR). In our analyses we carried out complete cases analyses (CCA) and to counter potential bias introduced by carrying out CCA we included relevant confounders (sex) which was related to missingness in our data [48].

## Results

### Sample description

Following quality control of the genetic data, a total of 8,654 children with genotyping data and at least one outcome measure were included in the analyses (Table 1). Information regarding age, sex, and ancestry/race for each timepoints can be found in Table S1. Our sensitivity analyses did not find evidence of sample overlap between our discovery (i.e., BMI GWAS; 25) and target cohort (i.e., ALSPAC) using LDSC, as the intercept from the genetic covariance (0.0068) was less than one standard error (0.0103) from zero, suggesting no sample overlap.

**Table 1:** Descriptive statistics of disordered eating behaviors and cognitions and BMI of the participants in the Avon Longitudinal Study of Parents and Children^a^

|  |  |  | Cases |  | Controls |  |
| --- | --- | --- | --- | --- | --- | --- |
| Binary outcomes | Age outcome measured (years) | Total N | N (%) | % Female | N | % Female |
| Fasting<sup>b</sup> | 14 | 4,584 | 300 (3.4%) | 83.7% | 4,284 | 53.3% |
| Fasting<sup>b</sup> | 16 | 3,844 | 516 (5.9%) | 90.7% | 3,328 | 53.8% |
| Fasting<sup>b</sup> | 18 | 2,586 | 143 (1.6%) | 93% | 2,443 | 62.1% |
| Binge eating<sup>b</sup> | 14 | 4,144 | 257 (2.9%) | 71.6% | 3,887 | 53.4% |
| Binge eating<sup>b</sup> | 16 | 3,336 | 434 (5.0%) | 83.2% | 2,902 | 55.3% |
| Binge eating<sup>b</sup> | 18 | 1,910 | 365 (4.2%) | 85.2% | 1,545 | 62.2% |
| Purging<sup>b</sup> | 16 | 3,871 | 237 (2.7%) | 92.4% | 3,634 | 56.7% |
| Purging<sup>b</sup> | 18 | 2,582 | 166 (1.9%) | 90.9% | 2,416 | 62.0% |
| Continuous outcomes | Age outcome measured (years) | Total N | mean | SD | Observed range (min) | Observed range (max) |
| Thin ideal internalization<sup>c</sup> | 14 | 4,496 | 15.33 | 2.69 | 5 | 25 |
| Body dissatisfaction<sup>d</sup> | 14 | 4,625 | 21.85 | 7.75 | 9 | 46.3 |
| Restrained eating<sup>e</sup> | 14 | 4,530 | 0.68 | 1.13 | 0 | 5 |
| Emotional eating<sup>e</sup> | 14 | 4,345 | 5.22 | 5.53 | 0 | 28 |
| External eating<sup>e</sup> | 14 | 3,995 | 8.45 | 3.34 | 0 | 21 |
| Age and sex adjusted body mass index (zBMI)<sup>f</sup> | 11 | 4,037 | 0.59 | 1.13 | -3.22 | 3.78 |
| **Total sample size<sup>g</sup>** |  | 8,654 |  |  |  |  |

SD = standard deviation.
<sup>a</sup>Full description of the Avon Longitudinal Study of Parents and Children is described elsewhere; [28–31].
<sup>b</sup>Assessed using questions modified from the Youth Risk Behavior Surveillance System questionnaire [32].
<sup>c</sup>Assessed the Ideal-Body Stereotype Scale-Revised with gender-specific items used to assess different aspects for boys and girls [6,33,34].
<sup>d</sup>Assessed using the Body Dissatisfaction scale [34,35].
<sup>e</sup>As measured by a modified version of the Dutch Eating Behavior Questionnaire [36].
<sup>f</sup>Calculated as weight in kilograms divided by height in meters squared. Height was measured to the nearest millimeter using a Harpenden Stadiometer (Holtain Ltd.) and weight was measured using the Tanita Body Fat analyzer (Tanita TBF UK Ltd.) to the nearest 50 grams. zBMI was calculated by standardising BMI by age and sex.
<sup>g</sup>Individuals with at least one outcome measurement (e.g., fasting, binge eating, body dissatisfaction) and available genotyping data.

### PGS analyses

#### BMI

In our confirmatory analyses we found that the BMI-PGS was significantly associated with BMI at age 11 years and 18 years and to zBMI measured at age 11 years (Table S2).

#### DE behaviors

The BMI-PGS was positively associated with fasting and binge eating at the ages 14, 16, and 18 years and with purging at age 16 years but not at age 18 years (Table 2). There was no significant interaction between the BMI-PGS and sex. When female participants were investigated separately we found similar effect sizes (Table S3). The effect of the best-fit BMI-PGS on fasting was fairly consistent; for one SD increase in the BMI-PGS children were 1.42 (95% CI 1.25 – 1.61), 1.29 (95% CI 1.17 – 1.42), and 1.26 (95% CI 1.06 – 1.51) times more likely to engage in fasting behavior at age 14, 16, and 18 years, respectively. We did not find evidence for a significant interaction between the PGS-BMI and fasting across the three ages indicating that the effect of the BMI-PGS is not age-dependent.

**Table 2:** Associations of body mass index polygenic score (BMI-PGS) with disordered eating behaviors and cognitions correcting for biological sex in the Avon Longitudinal Study of Parents and Children

| Binary outcomes | Age outcome measured (years) | Threshold<sup>a</sup> | N SNPs | R<sup>2</sup> | OR (95% CI)<sup>b</sup> | Q<sup>c</sup> |
| --- | --- | --- | --- | --- | --- | --- |
| Fasting | 14 | 0.085 | 33,379 | 0.021 | 1.42 (1.25-1.61) | <0.001 |
| Fasting | 16 | 0.17 | 43,956 | 0.012 | 1.29 (1.17-1.42) | <0.001 |
| Fasting | 18 | 0.1 | 36,088 | 0.008 | 1.26 (1.06-1.51) | 0.045 |
| Binge eating | 14 | 0.014 | 17,929 | 0.010 | 1.28 (1.12-1.46) | 0.003 |
| Binge eating | 16 | 0.0016 | 9,523 | 0.006 | 1.20 (1.08-1.34) | 0.006 |
| Binge eating | 18 | 0.0047 | 12,708 | 0.008 | 1.23 (1.09-1.39) | 0.008 |
| Purging | 16 | 0.00025 | 6,115 | 0.007 | 1.22 (1.07-1.41) | 0.02 |
| Purging | 18 | 0.033 | 23,731 | 0.005 | 1.20 (1.02-1.42) | 0.10 |
| Continuous outcomes | Age outcome measured (years) | Threshold<sup>a</sup> | N SNPs | R<sup>2</sup> | $\beta$ (95% CI)<sup>d</sup> | Q<sup>c</sup> |
| Thin ideal internalization | 14 | 0.5 | 66,077 | 0.003 | -0.15 (-0.23- -0.07) | 0.003 |
| Body dissatisfaction | 14 | 0.0013 | 9,075 | 0.015 | 0.99 (0.77-1.22) | <0.001 |
| Restrained eating | 14 | 0.1 | 36,088 | 0.015 | 0.14 (0.10-0.17) | <0.001 |
| Emotional eating | 14 | 0.0091 | 15,467 | 0.001 | 0.21 (0.052-0.38) | 0.046 |
| External eating | 14 | 0.0026 | 10,780 | 0.003 | -0.19 (-0.30- -0.09) | 0.003 |

SNP = single nucleotide polymorphism; R<sup>2</sup>, squared multiple correlation on the liability scale; OR, odds ratio; CI, confidence interval.
<sup>a</sup>The optimal *P*-value threshold for the inclusion of SNPs in the calculation of the body mass index (BMI) polygenic score (PGS) as determined by PRSice's high-resolution scoring [40].
<sup>b</sup>Odds ratio's reflect one standard deviation change in the standardized (to mean zero and standard deviation of one) BMI-PGS.
<sup>c</sup>Benjamini & Hochberg false discovery rate adjusting for the number of phenotypes tested [45].
<sup>d</sup>Standardized betas reflect one standard deviation increase in the standardized (to mean zero and standard deviation of one) BMI-PGS.

Children with higher BMI-PGS were also more likely to report binge eating at all three ages with ORs of 1.28 (95% CI 1.12 – 1.46), 1.20 (95% CI 1.08 – 1.34), and 1.23 (95% CI: 1.09 – 1.39), for the ages 14, 16, and 18 years, respectively. Furthermore, the effect of the BMI-PGS on binge eating did not differ across the three ages.

Individuals with higher BMI-PGS were 1.22 (95% CI 1.07 – 1.41) times more likely to report purging at age 16 years. We did not find an interaction between the BMI-PGS and age at reporting suggesting that the effect of the BMI-PGS did not differ across the reported ages (Table S4).

#### DE cognitions

The BMI-PGS was positively correlated with body dissatisfaction and with the DEBQ restrained and emotional scale (Table 2). The effect of this association was strongest for body dissatisfaction; children with a higher BMI-PGS (an increase of 1 SD were more likely to have higher scores on body dissatisfaction at age 14 years (Beta = 0.99, 95% CI 0.77 – 1.22). This effect was less pronounced for the restrained (Beta = 0.14, 95% CI: 0.10 – 1.17) and emotional eating (Beta = 0.21, 95% CI: 0.052 – 0.38) scale of the DEBQ in which a higher BMI-PGS corresponded with lower scores for both behaviors. In contrast to all other DE outcomes, we found that the BMI-PGS was negatively correlated with thin ideal internalization and with external eating; i.e., individuals with a higher BMI-PGS reported lower scores for thin ideal internalization (Beta = −0.15, 95% CI: −0.23 – −0.07) and external eating (Beta = −0.19, 95% CI: −0.30 – −0.09). There was no significant interaction between the BMI-PGS and sex in relation to the DE cognitions. Furthermore, the effect sizes of the association between the BMI-PGS and DE cognitions did not differ (95% confidence intervals overlapped) when female participants were analyzed separately (Table S3).

**Figure 2:**
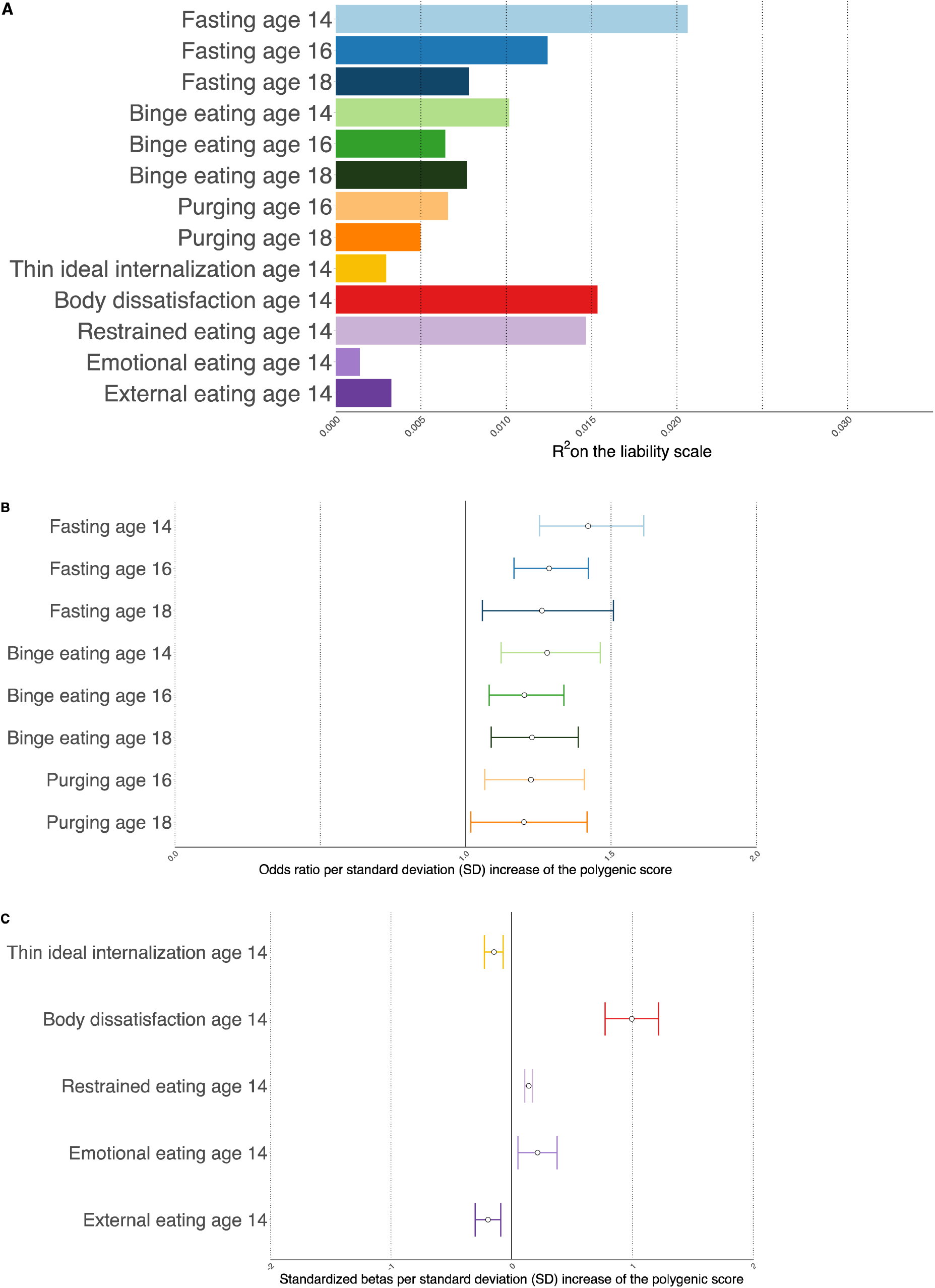
Association between the body mass index polygenic score (BMI-PGS) and disordered eating (DE) behaviors and cognitions in the Avon Longitudinal Study of Parents and Children [28–31]. Analyses were corrected for biological. Age is measured in years. **A:** Explained variance as measured by Nagelkerke’s Pseudo squared multiple correlation (R) for the DE phenotypes. All associations were statistically significant (False Discovery Rate-corrected Q-values < 0.05) except for purging at age 18 years. **B:** Effect size of the association between the standardized BMI-PGS (to mean zero and standard deviation of one) as measured by odds ratio’s (ORs) and the DE behaviors. The dots represent the point-estimates of the ORs for an increase of one standard deviation in the BM-PGS and the lines represent the 95% confidence interval of the point estimate. **C:** Effect size of the association between the standardized BMI-PGS (to mean zero and standard deviation of one) as measured by standardized beta’s and the DE cognitions. The dots represent the point-estimates of the standardized betas for an increase of one standard deviation in the BM-PGS and the lines represent the 95% confidence interval of the point estimate.

### Exploratory causal mediation analyses

Causal mediation analyses indicated that the zBMI scores mediated the association between the BMI-PGS and DE except for thin ideal internalization (Table 3). Estimate (β) of the average causal mediation effect ranged between −0.11 (external eating) and 0.93 (body dissatisfaction). For almost all mediation models, one standard deviation (SD) increase in the BMI-PGS corresponded with an increase in zBMI scores at age 11 years which in turn led to an increased probability of endorsing DE. The exception to this direction of effect was external eating; one standard deviation increase in the BMI-PGS led to an increase in zBMI scores at age 11 years, which then corresponded with lower external eating scores at 14 years.

**Table 3:** Exploratory causal mediation analysis of the association between the disordered eating outcomes and the standardized body mass index polygenic score (BMI-PGS) and the age sex adjusted body mass index z-scores at age 11 years as mediator. P-values for mediation were generated with bootstrapping methods.

| Phenotype | Age<sup>b</sup> | Total effect estimate<br>$\beta$ (95% CI) | Total effect<br>P-value | Average direct effect<br>$\beta$ (95% CI) | Average direct effect<br>P-value | Average causal mediation effect<br>$\beta$ (95% CI) | Average causal mediation<br>effect<br>P-value |
| --- | --- | --- | --- | --- | --- | --- | --- |
| Fasting | 14 | 0.022 (0.01-0.032) | <0.0001 | 0.011 (0.001-0.022) | 0.038 | 0.011 (0.007-0.014) | <0.0001 |
| Fasting | 16 | 0.035 (0.019-0.051) | <0.0001 | 0.021 (0.005-0.037) | 0.004 | 0.014 (0.008-0.019) | <0.0001 |
| Fasting | 18 | 0.014 (0.002-0.028) | 0.02 | 0.009 (-0.002-0.024) | 0.146 | 0.005 (0.001-0.01) | 0.03 |
| Binge eating | 14 | 0.01 (-0.001-0.02) | 0.07 | 0.004 (-0.007-0.014) | 0.464 | 0.006 (0.002-0.01) | <0.0001 |
| Binge eating | 16 | 0.015 (0.001-0.031) | 0.04 | -0.001 (-0.015-0.014) | 0.95 | 0.016 (0.01-0.022) | <0.0001 |
| Binge eating | 18 | 0.028 (0.006-0.053) | 0.01 | 0.01 (-0.013-0.034) | 0.348 | 0.017 (0.009-0.026) | <0.0001 |
| Purging | 16 | 0.011 (0.002-0.022) | 0.02 | 0.007 (-0.003-0.018) | 0.168 | 0.004 (0.001-0.008) | 0.006 |
| Purging | 18 | 0.014 (0.001-0.03) | 0.04 | 0.004 (-0.008-0.019) | 0.494 | 0.009 (0.005-0.014) | <0.0001 |
| Thin ideal internalization | 14 | -0.133 (-0.245--0.017) | 0.03 | -0.118 (-0.228--0.001) | 0.042 | -0.015 (-0.055-0.027) | 0.47 |
| Body dissatisfaction | 14 | 1.097 (0.819-1.378) | <0.0001 | 0.171 (-0.097-0.439) | 0.218 | 0.926 (0.798-1.061) | <0.0001 |
| Restrained eating | 14 | 0.161 (0.819-0.208) | <0.0001 | 0.03 (-0.014-0.072) | 0.162 | 0.13 (0.113-0.149) | <0.0001 |
| Emotional eating | 14 | 0.129 (-0.071-0.356) | 0.26 | 0.03 (-0.181-0.269) | 0.804 | 0.098 (0.023-0.176) | 0.01 |
| External eating | 14 | -0.195 (-0.335--0.053) | 0.006 | -0.087 (-0.228-0.06) | 0.212 | -0.108 (-0.162--0.057) | <0.0001 |

<sup>a</sup>The BMI-PGS was derived from summary statistics of the genome-wide association study (GWAS) carried out by the Genetic Investigation of Anthropometric Traits (GIANT) consortium [39] and were calculated for participants in the Avon Longitudinal Study of Parents and Children [28–31]. The age and sex adjusted BMI (zBMI) at age 11 years was included as a mediator in the causal mediation analyses that was carried out using the R package ‘mediation’ (version 4.4.6; 32) which is based on concepts proposed in modern causal inference [46]. Prior to the mediation analyses the BMI-PGS was standardized and the analysis was controlled for biological sex.
<sup>b</sup>Age was measured in years.
<sup>c</sup>Measured using the Dutch Eating Behavior Questionnaire (DEBQ; [36])

## Discussion

To our knowledge this is the largest study to date that has investigated whether a BMI-PGS is longitudinally associated with DE in a general population cohort (N = 8,654). We demonstrate that genetic factors that underlie BMI are also associated with DE suggesting a possible shared genetic etiology for BMI and DE. This association was mainly mediated through measured age and sex adjusted BMI (i.e., zBMI). Our results mirror findings from epidemiological studies that report higher BMI to be positively correlated with binge eating, emotional eating, restrained eating, purging, fasting, and body dissatisfaction, and negatively correlated with external eating [26,49–53]. The negative genetic correlation between the PGS-BMI and thin ideal internalization is also consistent with findings for AN, identified as negatively genetically correlated (rg = - 0.32) with BMI [23,24]. The non-significant association between the BMI-PGS and purging at age 18 years was likely due to the relatively rare endorsement of purging behavior (N = 166) emphasizing the need for larger samples. Furthermore, we did not find evidence for developmental differences; i.e., the strength of the association between the PGS-BMI and DE did not differ across ages.

It is important to emphasize that our study focused on DE as present in a general population cohort, which has important implications in interpreting our findings. Given the high incidence of overweight and obesity [54] in the general population, more individuals than ever could be at risk of developing DE and particularly those with elevated BMI-PGS could be at greater risk. Consistent with our findings, Nagata et al. reported that higher BMI-PGS is positively correlated with weight loss behaviors (e.g., fasting, use of laxatives) in a population cohort [25]. Findings from our group have also previously implicated BMI-related gene in adolescent binge eating in this sample [49]. Taken together, our data and those of others support the notion of a shared genetic etiology between DE and an elevated genetic propensity for higher BMI [25,49,55].

Results from our mediation analyses extend previous findings from Reed et al. that reported a causal effect of higher BMI in childhood on increased risk of DE at age 13 years using mendelian randomization [50]. The importance of BMI in predicting ED has also been highlighted in prospective studies in which BMI trajectories of children who develop an ED during adolescence were found to significantly deviate from children without an eating disorder [18]. We found differences in the estimates of the mediated effect by the age and sex corrected BMI (zBMI) in the association between the BMI-PGS and DE; this suggests that the effect of actual BMI in late childhood is important for body dissatisfaction, but less so for DE behaviors (e.g., purging and fasting for weight loss). As previously shown [3] cognitions such as body dissatisfaction are likely to be influenced by body image distortion and might hence be more impacted upon by environmental factors (e.g. comments and teasing about shape and weight). Furthermore, it is important to note that the zBMI scores did not mediate the association between the BMI-PGS and thin ideal internalization. The non-significant mediation for thin ideal internalization might suggest that other pathways apart from prior BMI might account for this association (e.g., environmental factors such as exposure to family or peer factors; [56]). Overall, our findings add to the considerable wealth of literature suggesting an important role for BMI in DE [3,18,23–25,50].

This study benefitted from a large discovery sample (N ∼ 789,224) that included participants from the GIANT consortium and the UK Biobank [39,57] and a relatively large target sample (N = 8,654). In addition, the availability of a wide range of DE outcomes collected at different time-points enriched our analyses. Especially the availability of objectively measured BMI at age 11 years allowed us to investigate the possible role BMI plays in the association between the PGS-BMI and DE.

Findings from this study should be interpreted in the context of some limitations. Participants were all recruited from the same geographical region in the south-west of England and therefore the results of this study might not be broadly generalizable to other populations. However, because of the homogeneity, this sample lends itself to genetic analyses as bias from population stratification is expected to be less pronounced [58]. It is important to note that ED symptoms included in this study were derived from self-report and the questions asked pertained to the previous year. This may have resulted in misclassification and recall bias. However, we would like to emphasize that observational measures on eating behaviors are not feasible in large cohort studies such as ALSPAC. We did not have information on DE behaviors and related cognitions in late childhood; occurrence of these behaviors prior to the measurement of BMI at age 11 years could have biased estimates of our mediation analysis. However, it is likely that prevalence of these behaviors and cognitions would have been very low in late childhood. We would also like to highlight that the focus of our study was on DE as present in the general population and while this information could improve our understanding of threshold ED, caution should be taken when interpreting these results in the context of threshold ED; a proportion of the individuals endorsing DE do not go on to develop an ED. Furthermore, considering the longitudinal nature of the study participants tend to drop out as time goes on leading to missing data. The missingness observed in our data was at random (MAR) and we tried to counter any bias resulting from our complete cases analyses by including covariates that were related to missingness in our dataset. We acknowledge that any remaining bias in our analyses could have biased our results towards the null [48].

The results from our mediation analyses are promising and suggest that joint approaches to the prevention of both obesity and DE might yield better clinical outcomes than those targeting the former or the latter independently [5]. Furthermore, increased awareness amongst pediatricians or general practitioners that children who have overweight or obesity are at increased risk of DE might aid in early screening, detection, and prevention of DE potentially mitigating the development of threshold ED [59]. BMI-PGS can also be used in conjunction with other PGS (e.g., eating disorder polygenic scores) and environmental risk factors in constructing clinical risk prediction models for ED; the clinical risk prediction model could for example aid in prediction and guide personalization of treatment, an approach that has shown to be fruitful in many somatic illnesses including coronary artery diseases [60]. However, we would like to note that at the current stage, given the effect sizes observed in this study, the utility of PGS for clinical application is premature.

In conclusion, our results suggest that genetic propensity for higher BMI is associated with DE, and that this association is mediated by actual measured BMI. Our findings add to the consistent epidemiological literature that implicates BMI in DE. The current study has demonstrated that DE and related cognitions needs to be understood in a broad context that includes anthropometry as well as behavioral and developmental components. Future research should focus on multifactorial risk and consider the role of environment as well as genetic predisposition to other related traits.

## Data Availability

Researchers can apply for the data at ALSPAC.

http://www.bristol.ac.uk/alspac/researchers/ourdata/

## Acknowledgments

This work was supported by the UK Medical Research Council and the Medical Research Foundation (ref: MR/R004803/1). The UK Medical Research Council and Wellcome (grant 102215/2/13/2) and the University of Bristol provide core support for ALSPAC. A comprehensive list of grants funding is available on the ALSPAC website (http://www.bristol.ac.uk/alspac/external/documents/grant-acknowledgements.pdf); This research was specifically funded by the National Institute of Health (MH087786-01 & MH115397-02) and the National Institute for Health Research (CS/01/2008/014). GWAS data was generated by Sample Logistics and Genotyping Facilities at Wellcome Sanger Institute and LabCorp (Laboratory Corporation of America) using support from 23andMe.

This publication is the work of the authors and MA will serve as guarantors for the contents of this paper.

CMB acknowledges funding from the Swedish Research Council (VR Dnr: 538-2013-8864). DSF works in a unit that receives funds from the University of Bristol and the UK Medical Research Council (MC_UU_00011/6). The funders were not involved in the design or conduct of the study; collection, management, analysis or interpretation of the data; or preparation, review or approval of the manuscript.

We wish to thank the Genetic Investigation of Anthropometric Traits (GIANT) consortium for providing the summary statistics of the GWAS on BMI. We are also extremely grateful to all the families who took part in the ALSPAC study, the midwives for their help in recruiting them, and the whole ALSPAC team, which includes interviewers, computer and laboratory technicians, clerical workers, research scientists, volunteers, managers, receptionists and nurses.

## Author Contributions

Conceptualization, Bianca De Stavola, Cynthia Bulik and Nadia Micali; Data curation, Moritz Herle and Nadia Micali; Formal analysis, Mohamed Abdulkadir, Moritz Herle, Bianca De Stavola and Nadia Micali; Funding acquisition, Nadia Micali; Investigation, Mohamed Abdulkadir, Moritz Herle, Bianca De Stavola and Nadia Micali; Project administration, Nadia Micali; Supervision, Bianca De Stavola, Ruth Loos, Rachel Bryant-Waugh, Cynthia Bulik and Nadia Micali; Visualization, Mohamed Abdulkadir; Writing – original draft, Mohamed Abdulkadir, Moritz Herle, Bianca De Stavola and Nadia Micali; Writing – review & editing, Mohamed Abdulkadir, Moritz Herle, Bianca De Stavola, Christopher Hübel, Diana Santos Ferreira, Ruth Loos, Rachel Bryant-Waugh, Cynthia Bulik and Nadia Micali.

## References

1. Swanson, S.A.; Crow, S.J.; Grange, D. Le; Swendsen, J.; Merikangas, K.R. Prevalence and Correlates of Eating Disorders in Adolescents. 2011, 68, 714–723.

2. Field, A.E.; Sonneville, K.R.; Micali, N.; Crosby, R.D.; Swanson, S.A.; Laird, N.M.; Treasure, J.; Solmi, F.; Horton, N.J. Prospective Association of Common Eating Disorders and Adverse Outcomes. Pediatrics 2012, 130, e289–e295.

3. Micali, N.; De Stavola, B.; Ploubidis, G.; Simonoff, E.; Treasure, J.; Field, A.E. Adolescent eating disorder behaviours and cognitions: Gender-specific effects of child, maternal and family risk factors. Br. J. Psychiatry 2015, 207, 320–327.

4. Solmi, F.; Sonneville, K.R.; Easter, A.; Horton, N.J.; Crosby, R.D.; Treasure, J.; Rodriguez, A.; Jarvelin, M.R.; Field, A.E.; Micali, N. Prevalence of purging at age 16 and associations with negative outcomes among girls in three community-based cohorts. J. Child Psychol. Psychiatry Allied Discip. 2015, 56, 87–96.

5. Neumark-Sztainer, D.; Wall, M.; Guo, J.; Story, M.; Haines, J.; Eisenberg, M. Obesity, disordered eating, and eating disorders in a longitudinal study of adolescents: How do dieters fare 5 years later? J. Am. Diet. Assoc. 2006, 106, 559–568.

6. Micali, N.; Solmi, F.; Horton, N.J.; Crosby, R.D.; Eddy, K.T.; Calzo, J.P.; Sonneville, K.R.; Swanson, S.A.; Field, A.E. Adolescent Eating Disorders Predict Psychiatric, High-Risk Behaviors and Weight Outcomes in Young Adulthood. J. Am. Acad. Child Adolesc. Psychiatry 2015, 54, 652–659.

7. Crow, S.; Eisenberg, M.E.; Story, M.; Neumark-Sztainer, D. Are body dissatisfaction, eating disturbance, and body mass index predictors of suicidal behavior in adolescents? A longitudinal study. J. Consult. Clin. Psychol. 2008, 76, 887–892.

8. Chamay-Weber, C.; Narring, F.; Michaud, P.-A. Partial eating disorders among adolescents: A review. J. Adolesc. Heal. 2005, 37, 416–426.

9. Berkman, N.; Bulik, C.; Brownley, K.; Lohr, K.; Sedway, J.; Rooks, A.; Gartlehner, G. Management of Eating Disorders. vidence Report/Technology Assessment No. 135. (Prepared by the RTI International-University of North Carolina Evidence-Based Practice Center under Contract No. 290-02-0016.) AHRQ Publication No. 06-E010. Rockville, MD: Age; 2006;

10. Keel, P.K.; Haedt, A. Evidence-Based Psychosocial Treatments for Eating Problems and Eating Disorders. J. Clin. Child Adolesc. Psychol. 2008, 37, 39–61.

11. Culbert, K.M.; Racine, S.E.; Klump, K.L. Research Review: What we have learned about the causes of eating disorders - a synthesis of sociocultural, psychological, and biological research. J. Child Psychol. Psychiatry 2015, 56, 1141–1164.

12. Culbert, K.M.; Racine, S.E.; Klump, K.L. The influence of gender and puberty on the heritability of disordered eating symptoms. In Behavioral neurobiology of eating disorders; Springer, 2010; pp. 177–185.

13. Martin, A.R.; Daly, M.J.; Robinson, E.B.; Hyman, S.E.; Neale, B.M. Predicting Polygenic Risk of Psychiatric Disorders. Biol. Psychiatry 2019, 86, 97–109.

14. Choi, S.W.; Shin, T.; Mak, H.; Reilly, P.F.O. A guide to performing Polygenic Risk Score analyses. bioRxiv 2018, 5, 11–13.

15. Tanofsky-Kraff, M.; Yanovski, S.Z.; Wilfley, D.E.; Marmarosh, C.; Morgan, C.M.; Yanovski, J.A. Eating-Disordered Behaviors, Body Fat, and Psychopathology in Overweight and Normal-Weight Children. J. Consult. Clin. Psychol. 2004, 72, 53–61.

16. Torstveit, M.K.; Aagedal-Mortensen, K.; Stea, T.H. More than half of high school students report disordered eating: A cross sectional study among Norwegian boys and girls. PLoS One 2015, 10, 1–15.

17. Flament, M.F.; Henderson, K.; Buchholz, A.; Obeid, N.; Nguyen, H.N.T.; Birmingham, M.; Goldfield, G. Weight Status and DSM-5 Diagnoses of Eating Disorders in Adolescents From the Community. J. Am. Acad. Child Adolesc. Psychiatry 2015, 54, 403-411.e2.

18. Yilmaz, Z.; Gottfredson, N.C.; Zerwas, S.C.; Bulik, C.M.; Micali, N. Developmental Premorbid Body Mass Index Trajectories of Adolescents With Eating Disorders in a Longitudinal Population Cohort. J. Am. Acad. Child Adolesc. Psychiatry 2019, 58, 191–199.

19. Herle, M.; Stavola, B. De; Hübel, C.; Abdulkadir, M.; Ferreira, D.S.; Loos, R.J.F.; Bryant-Waugh, R.; Bulik, C.M.; Micali, N. A longitudinal study of eating behaviours in childhood and later eating disorder behaviours and diagnoses. Br. J. Psychiatry 2019, 1–7.

20. Föcker, M.; Bühren, K.; Timmesfeld, N.; Dempfle, A.; Knoll, S.; Schwarte, R.; Egberts, K.M.; Pfeiffer, E.; Fleischhaker, C.; Wewetzer, C.; et al. The relationship between premorbid body weight and weight at referral, at discharge and at 1-year follow-up in anorexia nervosa. Eur. Child Adolesc. Psychiatry 2015, 24, 537–544.

21. Albuquerque, D.; Stice, E.; Rodríguez, R.; Licíno, L.-R.; Manco, L.; Nóbrega, C. Current review of genetics of human obesitylll: from molecular mechanisms to an evolutionary perspective. Mol. Genet. Genomics 2015, 1191–1221.

22. Khera, A. V.; Chaffin, M.; Wade, K.H.; Zahid, S.; Brancale, J.; Xia, R.; Distefano, M.; Senol-Cosar, O.; Haas, M.E.; Bick, A.; et al. Polygenic Prediction of Weight and Obesity Trajectories from Birth to Adulthood. Cell 2019, 177, 587-596.e9.

23. Watson, H.J.; Yilmaz, Z.; Thornton, L.M.; Hübel, C.; Coleman, J.R.I.I.; Gaspar, H.A.; Bryois, J.; Hinney, A.; Leppä, V.M.; Mattheisen, M.; et al. Genome-wide association study identifies eight risk loci and implicates metabo-psychiatric origins for anorexia nervosa. Nat. Genet. 2019, 51, 1207–1214.

24. Duncan, L.; Yilmaz, Z.; Gaspar, H.; Walters, R.; Goldstein, J.; Anttila, V.; Bulik-Sullivan, B.; Ripke, S.; Thornton, L.; Hinney, A.; et al. Significant locus and metabolic genetic correlations revealed in genome-wide association study of anorexia nervosa. Am. J. Psychiatry 2017, 174, 850–858.

25. Nagata, J.M.; Braudt, D.B.; Domingue, B.W.; Bibbins-Domingo, K.; Garber, A.K.; Griffiths, S.; Murray, S.B. Genetic risk, body mass index, and weight control behaviors: Unlocking the triad. Int. J. Eat. Disord. 2019, 52, 825–833.

26. Snoek, H.M.; Engels, R.C.M.E.; van Strien, T.; Otten, R. Emotional, external and restrained eating behaviour and BMI trajectories in adolescence. Appetite 2013, 67, 81–87.

27. Juarascio, A.S.; Forman, E.M..; Timko, C.A.; Herbert, J.D.; Butryn, M.; Lowe, M. Implicit internalization of the thin ideal as a predictor of increases in weight, body dissatisfaction, and disordered eating. Eat. Behav. 2011, 12, 207–213.

28. Fraser, A.; Macdonald-wallis, C.; Tilling, K.; Boyd, A.; Golding, J.; Davey smith, G.; Henderson, J.; Macleod, J.; Molloy, L.; Ness, A.; et al. Cohort profile: The avon longitudinal study of parents and children: ALSPAC mothers cohort. Int. J. Epidemiol. 2013, 42, 97–110.

29. Golding; Pembrey Jones; The Alspac Study Team ALSPAC-The Avon Longitudinal Study of Parents and Children. Paediatr. Perinat. Epidemiol. 2001, 15, 74–87.

30. Golding, J.; Team, S. The Avon Longitudinal Study of Parents and Children (ALSPAC) – study design and collaborative opportunities. 2004, 119–123.

31. Boyd, A.; Golding, J.; Macleod, J.; Lawlor, D.A.; Fraser, A.; Henderson, J.; Molloy, L.; Ness, A.; Ring, S.; Smith, G.D. Cohort profile: The ‘Children of the 90s’-The index offspring of the avon longitudinal study of parents and children. Int. J. Epidemiol. 2013, 42, 111–127.

32. Kann, L.; Warren, C.W.; Harris, W.A.; Collins, J.L.; Williams, B.I.; Ross, J.G.; Kolbe, L.J. Youth Risk Behavior Surveillance-United States, 1995. J. Sch. Health 1996, 66, 365–377.

33. Stice, E. Modeling of eating pathology and social reinforcement of the thin-ideal predict onset of bulimic symptoms. Behav. Res. Ther. 1998, 36, 931–944.

34. Calzo, J.P.; Austin, S.B.; Micali, N. Sexual orientation disparities in eating disorder symptoms among adolescent boys and girls in the UK. Eur. Child Adolesc. Psychiatry 2018, 27, 1–8.

35. Stice, E. A prospective test of the dual-pathway model of bulimic pathology: Mediating effects of dieting and negative affect. J. Abnorm. Psychol. 2001, 110, 124–135.

36. van Strien, T.; Frijters, J.E.R.; Bergers, G.P.A.; Defares, P.B. The Dutch Eating Behavior Questionnaire (DEBQ) for assessment of restrained, emotional, and external eating behavior. Int. J. Eat. Disord. 1986, 5, 295–315.

37. Schaumberg, K.; Zerwas, S.; Goodman, E.; Yilmaz, Z.; Bulik, C.M.; Micali, N. Anxiety disorder symptoms at age 10 predict eating disorder symptoms and diagnoses in adolescence. J. Child Psychol. Psychiatry Allied Discip. 2018.

38. Cole, T.J.; Bellizzi, M.; Flegal, K.; Dietz, W. Establishing a standard definition for child overweight and obesity worldwide: international survey. BMJ 2000, 320, 1240–1240.

39. Yengo, L.; Sidorenko, J.; Kemper, K.E.; Zheng, Z.; Wood, A.R.; Weedon, M.N.; Frayling, T.M.; Hirschhorn, J.; Yang, J.; Visscher, P.M. Meta-analysis of genome-wide association studies for height and body mass index in ~700000 individuals of European ancestry. Hum. Mol. Genet. 2018, 27, 3641–3649.

40. Euesden, J.; Lewis, C.M.; O’Reilly, P.F. PRSice: Polygenic Risk Score software. Bioinformatics 2015, 31, 1466–1468.

41. Choi, S.W.; O’Reilly, P.F. PRSice-2: Polygenic Risk Score software for biobank-scale data. Gigascience 2019, 8, 1–6.

42. Purcell, S.; Neale, B.; Todd-Brown, K.; Thomas, L.; Ferreira, M.A.R.; Bender, D.; Maller, J.; Sklar, P.; de Bakker, P.I.W.; Daly, M.J.; et al. PLINK: a tool set for whole-genome association and population-based linkage analyses. Am. J. Hum. Genet. 2007, 81, 559–75.

43. Bulik-Sullivan, B.; Loh, P.R.; Finucane, H.K.; Ripke, S.; Yang, J.; Patterson, N.; Daly, M.J.; Price, A.L.; Neale, B.M.; Corvin, A.; et al. LD score regression distinguishes confounding from polygenicity in genome-wide association studies. Nat. Genet. 2015, 47, 291–295.

44. Lee, S.H.; Goddard, M.E.; Wray, N.R.; Visscher, P.M. A better coefficient of determination for genetic profile analysis. Genet. Epidemiol. 2012, 36, 214–224.

45. Benjamini, Y.; Hochberg, Y. Controlling the False Discovery Ratelll: a Practical and Powerful Approach to Multiple Testing. J. R. Stat. Soc. Ser. B 1995, 57, 289–300.

46. VanderWeele, T. Explanation in causal inference: methods for mediation and interaction; Oxford University Press, 2015;

47. Tingley, D.; Yamamoto, T.; Hirose, K.; Keele, L.; Imai, K. mediationlll: R Package for Causal Mediation Analysis. J. Stat. Softw. 2014, 59, 1–38.

48. White, I.R.; Carlin, J.B. Bias and efficiency of multiple imputation compared with complete-case analysis for missing covariate values. Stat. Med. 2010, 29, 2920–2931.

49. Micali, N.; Field, A.E.; Treasure, J.L.; Evans, D.M. Are obesity risk genes associated with binge eating in adolescence? Obesity 2015, 23, 1729–1736.

50. Reed, Z.E.; Micali, N.; Bulik, C.M.; Davey Smith, G.; Wade, K.H. Assessing the causal role of adiposity on disordered eating in childhood, adolescence, and adulthood: a Mendelian randomization analysis. Am. J. Clin. Nutr. 2017, ajcn154104.

51. Bucchianeri, M.M.; Arikian, A.J.; Hannan, P.J.; Eisenberg, M.E.; Neumark-Sztainer, D. Body dissatisfaction from adolescence to young adulthood: Findings from a 10-year longitudinal study. Body Image 2013, 10, 1–7.

52. Keel, P.K.; Baxter, M.G.; Heatherton, T.F.; Joiner, T.E. A 20-year longitudinal study of body weight, dieting, and eating disorder symptoms. J. Abnorm. Psychol. 2007, 116, 422–432.

53. Stice, E.; Gau, J.M.; Rohde, P.; Shaw, H. Risk factors that predict future onset of each DSM–5 eating disorder: Predictive specificity in high-risk adolescent females. J. Abnorm. Psychol. 2017, 126, 38–51.

54. Bentham, J.; Di Cesare, M.; Bilano, V.; Bixby, H.; Zhou, B.; Stevens, G.A.; Riley, L.M.; Taddei, C.; Hajifathalian, K.; Lu, Y.; et al. Worldwide trends in body-mass index, underweight, overweight, and obesity from 1975 to 2016: a pooled analysis of 2416 population-based measurement studies in 128·9 million children, adolescents, and adults. Lancet 2017, 390, 2627–2642.

55. Hinney, A.; Kesselmeier, M.; Jall, S.; Volckmar, A.-L.; Föcker, M.; Antel, J.; Heid, I.M.; Winkler, T.W.; Grant, S.F.A.; Guo, Y.; et al. Evidence for three genetic loci involved in both anorexia nervosa risk and variation of body mass index. Mol. Psychiatry 2017, 22, 192–201.

56. Field, A.E.; Javaras, K.M.; Aneja, P.; Kitos, N.; Camargo, C.A.; Taylor, C.B.; Laird, N.M. Family, Peer, and Media Predictors of Becoming Eating Disordered. Arch. Pediatr. Adolesc. Med. 2008, 162, 574.

57. Bycroft, C.; Freeman, C.; Petkova, D.; Band, G.; Elliott, L.T.; Sharp, K.; Motyer, A.; Vukcevic, D.; Delaneau, O.; O’Connell, J.; et al. The UK Biobank resource with deep phenotyping and genomic data. Nature 2018, 562, 203–209.

58. Hellwege, J.N.; Keaton, J.M.; Giri, A.; Gao, X.; Velez Edwards, D.R.; Edwards, T.L. Population Stratification in Genetic Association Studies. Curr. Protoc. Hum. Genet. 2017, 95, 1.22.1-1.22.23.

59. Hayes, J.F.; Fitzsimmons-Craft, E.E.; Karam, A.M.; Jakubiak, J.; Brown, M.L.; Wilfley, D.E. Disordered Eating Attitudes and Behaviors in Youth with Overweight and Obesity: Implications for Treatment. Curr. Obes. Rep. 2018, 7, 235–246.

60. Lambert, S.A.; Abraham, G.; Inouye, M. Towards clinical utility of polygenic risk scores. Hum. Mol. Genet. 2019, 00, 1–10.

